# Examining Risk Factors Related to Cardiac Rehabilitation Cessation Among Advanced Heart Failure Patients

**DOI:** 10.1101/2023.11.22.23298936

**Authors:** Sharnendra K. Sidhu, Bernard S. Kadosh, François Haas, Ying Tang, Greg Sweeney, Alicia Pierre, Jonathan Whiteson, Edward Katz, Alex Reyentovich, John A. Dodson

## Abstract

**Background:** Cardiac rehabilitation (CR) has shown benefits in heart transplant and left ventricular assist device (LVAD) recipients, but variable patterns of attendance remain poorly understood. We therefore aimed to describe CR adherence and factors associated with cessation in this population.

**Methods:** We performed a retrospective review of heart transplant and LVAD recipients who attended at least one CR session at a tertiary medical center (2013-2022). Complete adherence was defined as attending all 36 CR sessions. Primary reasons for cessation prior to 36 sessions were recorded. We compared post-operative complications and length of stay (following heart transplant or LVAD) between participants with and without complete adherence using logistic regression. In a subgroup analysis of participants with complete adherence, we compared changes in METs, exercise time, and peak VO_2_ using paired-sample t tests.

**Results:** There were 137 heart transplant and LVAD recipients (median age 56.9 years, 73.7% male) who attended CR. Ninety-one percent either completed all 36 CR sessions or fewer than 24 sessions. Among those without complete adherence (n=74), 71.6% reported medical reasons and 15.0% reported personal reasons for cessation. Participants without complete adherence experienced more post-operative complications (43.9% vs 34.4%, p = 0.02) and major bleeding (22.7% vs 15.2%, p = 0.02) prior to enrolling in CR, but there were no other significant differences between groups. Participants with complete adherence experienced significant improvements in exercise time (160.9 seconds), METs (0.8), and peak VO_2_ (1.5 mL/kg per min).

**Conclusions:** Among heart transplant and LVAD recipients, there was a bimodal distribution of CR attendance; nearly half completed all 36 sessions. Among those with complete adherence, there were significant improvements in exercise measures, underscoring the important benefits of CR in this population.

## Introduction

Heart transplantation and left ventricular assist devices (LVADs) are effective in end stage heart failure (HF) refractory to medical therapy.^1^ Despite the effectiveness of these therapies in prolonging life, recipients are often deconditioned and their exercise capacity remains severely limited.^2–3^ Reduced exercise tolerance is associated with worse survival in both populations,^4–5^ and strategies to improve engagement in exercise therefore represents a potential opportunity to mitigate this risk. Cardiac rehabilitation (CR), which provides structured exercise and lifestyle modification under direct medical supervision, has been significantly associated with improvements in many measures of exercise capacity following heart transplantation and LVAD implantation.^6–11^

Many published analyses suggest a dose-response relationship correlating a greater number of CR sessions attended with lower risk of major cardiovascular events.^12–15^ While studies have demonstrated that CR has benefits in heart transplant^16–17^ and LVAD recipients^18–19^, there are still multiple barriers that include lack of referral to CR, failure to attend the first visit (even if referred), and attrition (failure to attend all 36 recommended sessions).^20–24^ While lack of referral may be due to clinical inertia and can be improved, for example, through systems-based solutions such as electronic health record (EHR) prompts^25^, attrition remains a problem that is poorly understood. However, it is known that broadly, among all CR eligible Medicare beneficiaries in 2016, only 27% of those who initiated CR completed a full course.^23^

In this context, our aims were to describe patterns of CR termination, and its predictors, among an advanced heart failure population that included heart transplant or LVAD recipients. These individuals remain exceedingly debilitated following surgery and have a propensity for both prolonged hospital stays and consequential post-operative complications (i.e. infection, bleeding, primary device or graft failure), which may contribute to greater rates of attrition compared with other populations.

## Methods

This retrospective study included all patients who received a heart transplant or LVAD and subsequently participated in at least 1 CR session at New York University (NYU) Langone Rusk Rehabilitation from January 2013 through December 2022. Patients were referred to CR by a cardiologist or other healthcare provider as part of standard care. This study was approved as a retrospective chart review by the institutional review board of NYU Langone Health; informed consent was not required.

Demographic information, comorbidities, biometrics [weight (kg), height (cm), body mass index (BMI; kg/m^2^), creatinine (mg/dL)], and cardiomyopathy subclass (ischemic vs. nonischemic) were extracted for each participant from the EHR. Comorbidities included history of arrhythmia, coronary artery disease, diabetes, hypertension, hyperlipidemia, pulmonary disease, obesity, depression, prior stroke, and smoking history. BMI was abstracted from the CR intake visit, with obesity defined as BMI >30 kg/m^2^. Creatine clearance was calculated using the laboratory measurement closest to hospital discharge. Data on duration of hospitalization and adverse post-operative events (defined as major bleeding event, infection, need for dialysis, need for reoperation) were also extracted from the EHR.

We recorded the number of CR sessions attended and CR completion status (with complete adherence defined as participation in all 36 CR sessions). Participants were divided into groups based on the number of CR sessions they completed (1 to 11, 12 to 23, 24 to 35, or 36 sessions). As part of routine CR care, participants underwent exercise stress testing by either Bruce or modified Bruce protocol and cardiopulmonary exercise testing before and, if they completed all 36 sessions, again after CR. Metabolic equivalent tasks (MET) and exercise time were assessed by exercise stress test, whereas peak VO_2_ was assessed by cardiopulmonary exercise test. Not all individuals were able to complete post-CR functional testing despite completion of 36 CR sessions due to patient-specific factors or logistical reasons. Patient-cited reasons for cessation from CR prior to 36 sessions were elicited by rehabilitation staff and categorized as: medical, personal, insurance expense, COVID-19 pandemic, or called for heart transplant (LVAD patients).

### Statistical Analysis

Baseline characteristics, duration of hospitalization (following heart transplant or LVAD implantation), and post-operative complications were compared among participants who completed all 36 CR sessions versus those of participants who did not. We used 2-sample t-tests for parametric continuous variables, Kruskal-Wallis tests for nonparametric continuous variables, and Fisher exact tests for categorical variables. Given limited sample sizes, race was categorized as White versus non-White (consisting of individuals who self-identified as Black, Asian, or “Other” on clinical documentation). Multivariable logistic regressions were employed to adjust for age, sex, and diabetes when determining the association of both hospitalization duration and post-operative complications with cessation from CR prior to 36 sessions. For participants who completed all 36 CR sessions and had both pre– and post–exercise stress test and/or cardiopulmonary exercise test, we performed a subgroup analysis to evaluate changes in METs, exercise time, and peak VO_2_ before and after CR using paired-sample t-tests. The threshold for statistical significance was set at a two-tailed p value of 0.05. All statistical analyses were performed using R Studio (R Studio version 4.0.3).

## Results

Our sample included 137 heart transplant and LVAD recipients (heart transplant = 62, LVAD = 75) who attended at least 1 CR session from January 2013 through December 2022. Median age was 56.9 years (IQR 47.9-62.4) and 73.7% were male (Table 1). Self-identified race of patients was 34.3% Black, 26.3% White, 5.8% Asian, 32.1% Other, and 1.5% unknown. Hispanic ethnicity was self-reported by 21.2% of patients. The majority of patients had nonischemic cardiomyopathy (66.4%, vs. 30.6% with ischemic cardiomyopathy). Common comorbidities included hypertension (68.4%), hyperlipidemia (66.2%), and current or former smoking history (46.7%). Slightly over half of patients (51.1%) received their heart transplant or LVAD within the NYU system, with the remaining undergoing their operation elsewhere.

**Table 1:** Baseline Characteristics of Heart Transplant and LVAD Patients Participating in CR and Those Completing the Program.

| Characteristic | Did not complete CR (n=74) | Completed CR (n=63) | p-value |
| --- | --- | --- | --- |
| **Demographics** |  |  |  |
| Age, y | 56.0 [48.1, 62.3] | 57.0 [47.9, 62.1] | 0.628 |
| Male sex | 54 (73.0%) | 47 (74.6%) | 0.848 |
| White race | 18 (24.3%) | 18 (28.6%) | 0.697 |
| Non-White race† | 56 (75.7%) | 43 (68.3%) | 0.346 |
| Hispanic ethnicity | 13 (17.6%) | 16 (25.4%) | 0.298 |
| Medicare | 21 (28.4%) | 27 (43.5%) | 0.074 |
| Medicaid | 35 (47.3%) | 27 (43.5%) | 0.731 |
| BMI, kg/m<sup>2</sup> | 28.03 (6.6) | 26.73 (5.4) | 0.222 |
| CrCl at discharge, mL/min | 83.80 [63.6, 104.0] | 87.0 [64.0, 107.6] | 0.648 |
| **Comorbidities** |  |  |  |
| Coronary artery disease | 36 (48.6%) | 23 (37.1%) | 0.224 |
| Diabetes | 37 (50.0%) | 20 (32.3%) | 0.055 |
| Hypertension | 54 (73.0%) | 39 (62.9%) | 0.267 |
| Hyperlipidemia | 50 (67.6%) | 40 (64.5%) | 0.720 |
| Obesity | 30 (40.5%) | 18 (29.0%) | 0.208 |
| Depression | 13 (17.6%) | 12 (19.4%) | 0.827 |
| Stroke | 19 (25.7%) | 16 (25.8%) | 1.000 |
| COPD | 12 (16.2%) | 8 (12.9%) | 0.634 |
| Atrial Fibrillation | 30 (40.5%) | 24 (38.7%) | 0.862 |
| Current/former smoker | 35 (47.3%) | 29 (46.8%) | 0.865 |
| Data are given as mean (SD) for parametric continuous variables and median (IQR) for nonparametric continuous variables. Categorical variables are given as numbers (percentage). 2-sample tests for parametric continuous variables, Kruskal Wallis tests for nonparametric continuous variables, and Fisher exact tests for categorical variables. BMI, body mass index; CR, cardiac rehabilitation; COPD, chronic obstructive pulmonary disease; CrCl, creatinine clearance; LVAD: left ventricular assist device.<br>†Non-White race included individuals self-identifying as Asian, Black, or Other on clinical documentation. |  |  |  |

On average, patients completed an average of 24.6 CR sessions (SD 12.4). Sixty-three (46%) had complete adherence to all 36 sessions (heart transplant: 44.0%, LVAD: 48.3%; Figures 1 and 2). Among remaining patients, the mean number of sessions attended was 14.9 (SD 9.0), with 31 attending 1-11 sessions, 31 attending 12-23 sessions, and 12 attending 24-35 sessions. There was a bimodal distribution of attendance; 91% of patients either completed all 36 CR sessions or completed fewer than 24 CR sessions. Among those who did not complete all 36 CR sessions, 71.6% reported medical reasons for cessation, 15.0% reported personal reasons, 5.4% reported concerns related to the COVID-19 pandemic, 2.7% reported out-of-pocket costs, 2.7% were called for transplant, and 2.7% did not report a reason (Figure 3).

**Figure 1.**
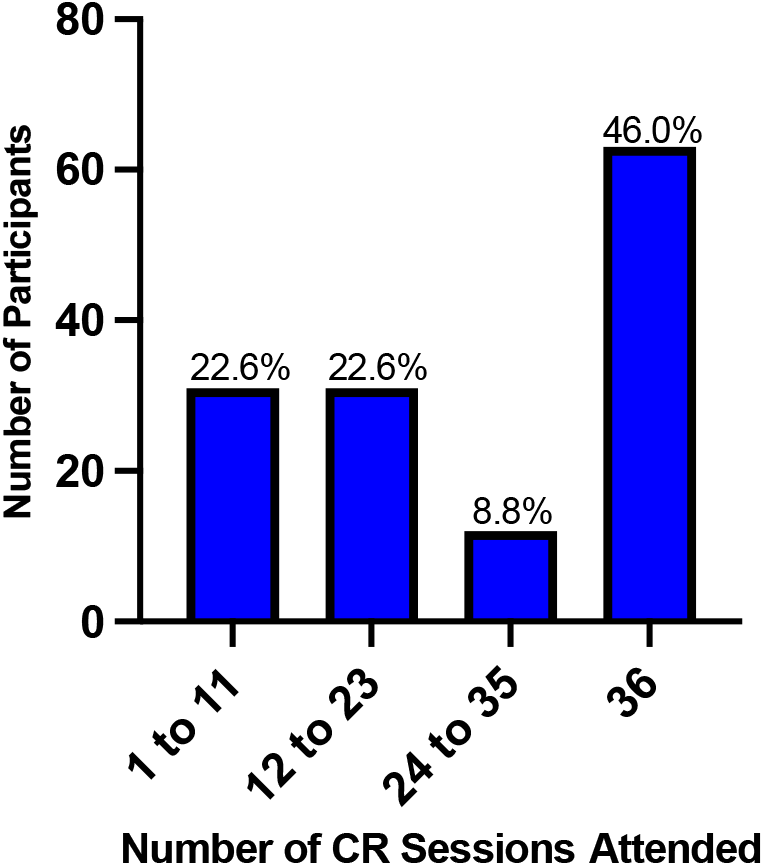
Distribution of heart transplant and left ventricular assist device recipients by number of cardiac rehabilitation sessions attended.

**Figure 2.**
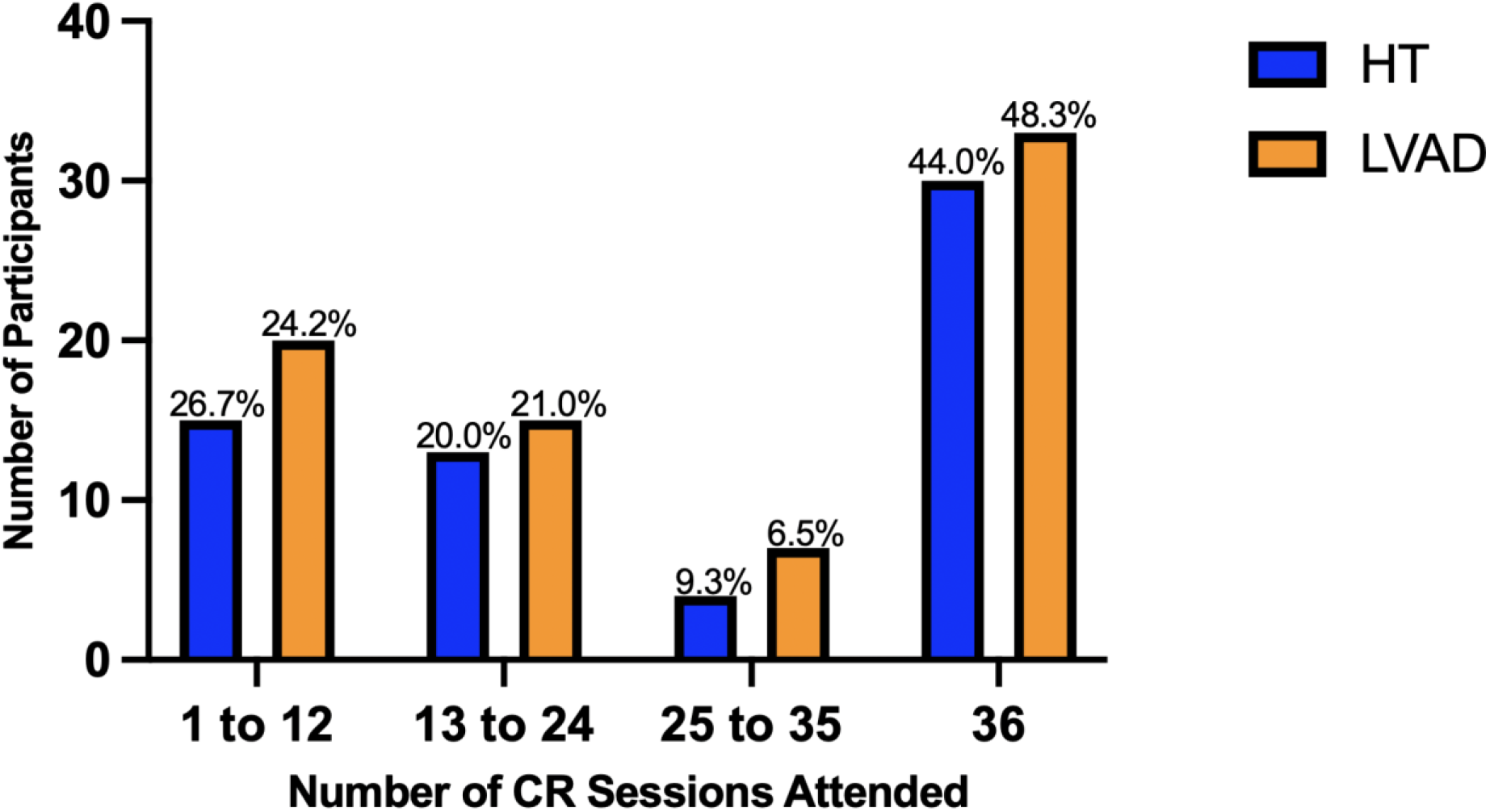
Subgroup distribution of heart transplant and left ventricular assist device recipients by number of cardiac rehabilitation sessions attended.

**Figure 3.**
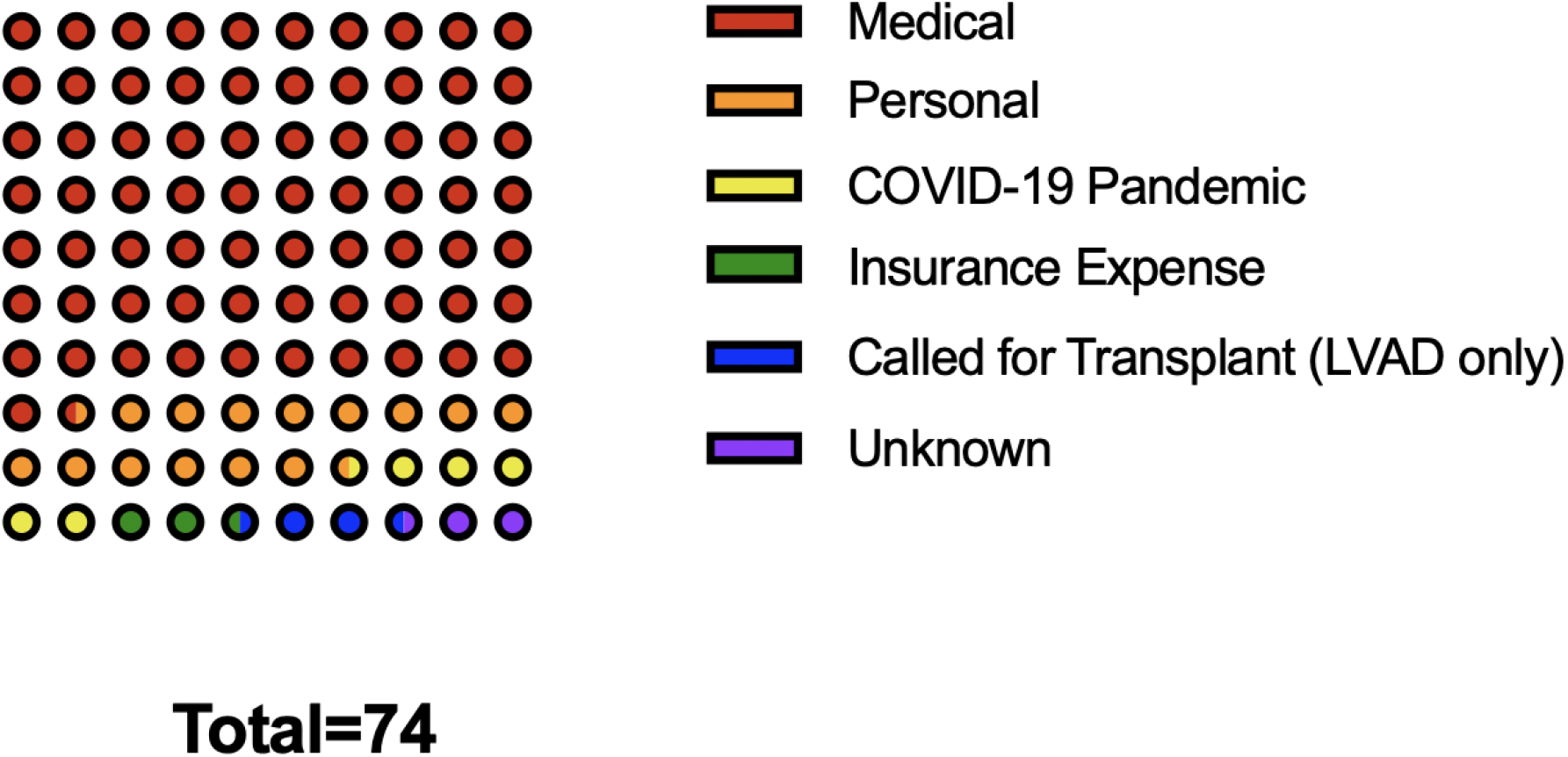
10×10 dot plot depicting patient cited reasons for cessation among those who did not complete all 36 cardiac rehabilitation sessions.

Patient age, sex, and comorbidities were not associated with CR completion. However, patients who did not complete CR experienced more overall post-operative complications (43.9% vs 34.4%, p = 0.02) as well as post-operative major bleeding (22.7% vs 15.2%, p = 0.02) at the time of index hospitalization for heart transplantation or LVAD placement compared with those who completed CR, including after multivariable adjustment (Table 2). There was no significant association between CR completion and other factors including duration of hospitalization, need for reoperation, infection, or need for dialysis.

**Table 2:**
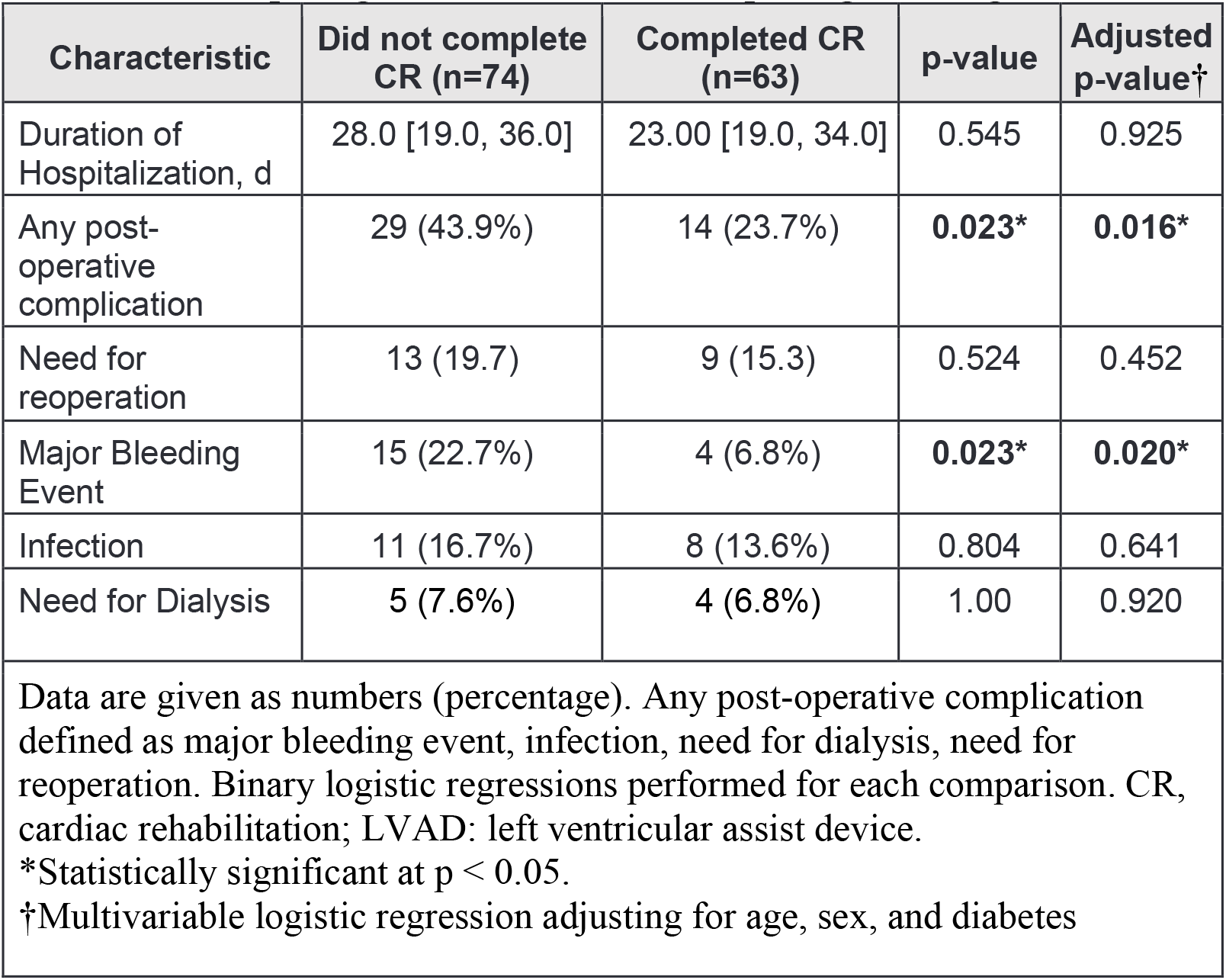
Post-Operative Period and Complications of Heart Transplant and LVAD Patients Participating in CR and Those Completing the Program.

| Characteristic | Did not complete CR (n=74) | Completed CR (n=63) | p-value | Adjusted p-value† |
| --- | --- | --- | --- | --- |
| Duration of Hospitalization, d | 28.0 [19.0, 36.0] | 23.00 [19.0, 34.0] | 0.545 | 0.925 |
| Any post-operative complication | 29 (43.9%) | 14 (23.7%) | **0.023\*** | **0.016\*** |
| Need for reoperation | 13 (19.7) | 9 (15.3) | 0.524 | 0.452 |
| Major Bleeding Event | 15 (22.7%) | 4 (6.8%) | **0.023\*** | **0.020\*** |
| Infection | 11 (16.7%) | 8 (13.6%) | 0.804 | 0.641 |
| Need for Dialysis | 5 (7.6%) | 4 (6.8%) | 1.00 | 0.920 |
| Data are given as numbers (percentage). Any post-operative complication defined as major bleeding event, infection, need for dialysis, need for reoperation. Binary logistic regressions performed for each comparison. CR, cardiac rehabilitation; LVAD: left ventricular assist device.<br>*Statistically significant at $p < 0.05$.<br>†Multivariable logistic regression adjusting for age, sex, and diabetes |  |  |  |  |

### Subgroup Analysis

Among the 63 patients who completed CR, pre- and post-CR exercise test results (exercise stress test and/or cardiopulmonary exercise test) were available for 49 (77.8%). On average, they experienced significant improvements in exercise time by 160.9 seconds (95% CI, 119.5-202.4, baseline 415.9 seconds, p<0.0001), METs by 0.8 (95% CI 0.3-1.2, baseline 4.0, p<0.001), and peak VO_2_ by 1.5 mL/kg per min (95% CI 0.1-2.8, baseline 15.3 mL/kg per min, p=0.03; Table 3 and Figure 4).

**Table 3:**
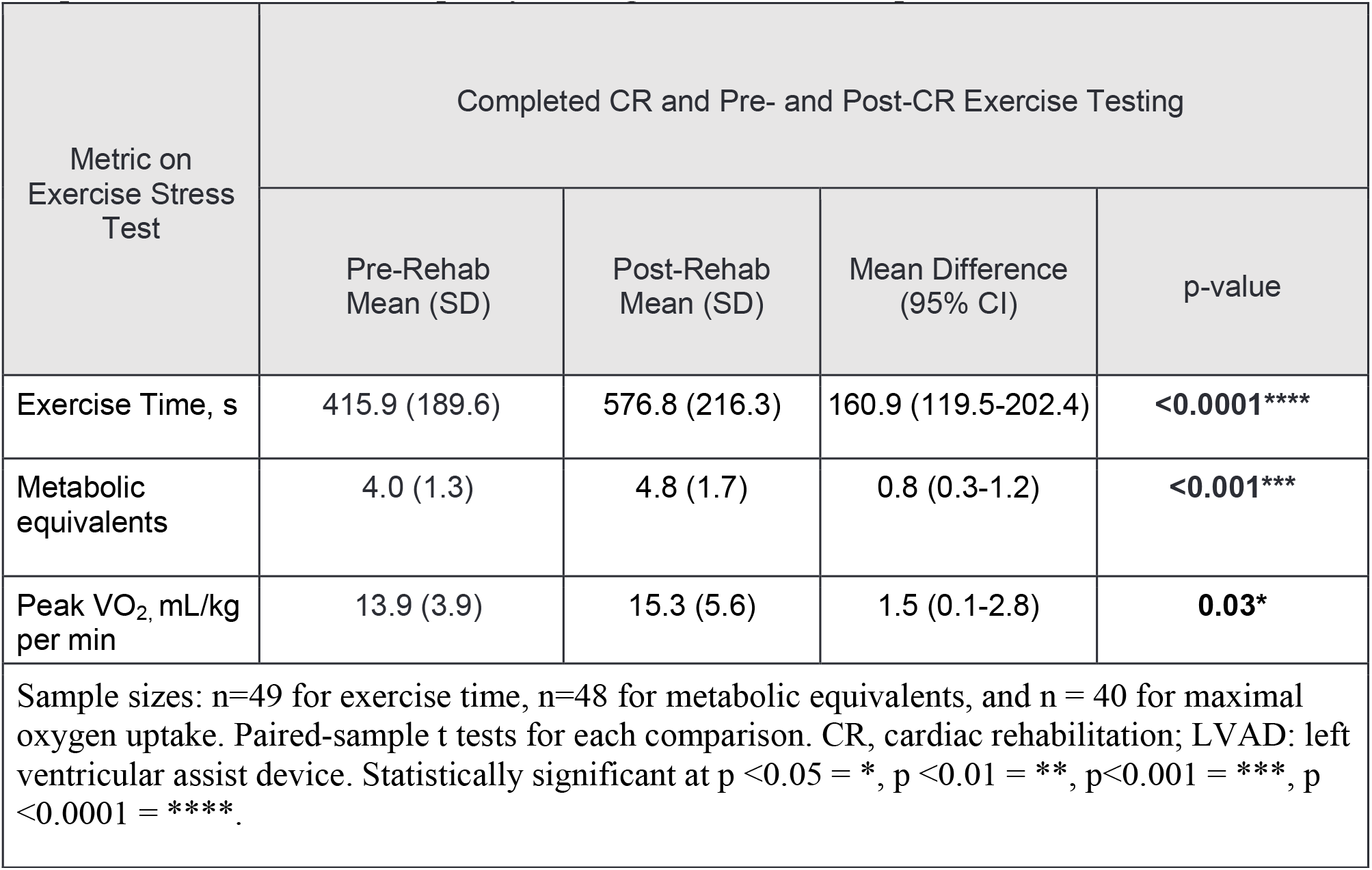
Baseline Exercise Capacity of Heart Transplant and LVAD Patients and Improvement in Exercise Capacity Among Those Who Completed CR.

| Metric on Exercise Stress Test | Completed CR and Pre- and Post-CR Exercise Testing |  |  |  |
| --- | --- | --- | --- | --- |
|  | Pre-Rehab Mean (SD) | Post-Rehab Mean (SD) | Mean Difference (95% CI) | p-value |
| Exercise Time, s | 415.9 (189.6) | 576.8 (216.3) | 160.9 (119.5-202.4) | **<0.0001\*\*\*\*** |
| Metabolic equivalents | 4.0 (1.3) | 4.8 (1.7) | 0.8 (0.3-1.2) | **<0.001\*\*\*** |
| Peak VO<sub>2</sub>, mL/kg per min | 13.9 (3.9) | 15.3 (5.6) | 1.5 (0.1-2.8) | **0.03\*** |
| Sample sizes: n=49 for exercise time, n=48 for metabolic equivalents, and n = 40 for maximal oxygen uptake. Paired-sample t tests for each comparison. CR, cardiac rehabilitation; LVAD: left ventricular assist device. Statistically significant at p <0.05 = *, p <0.01 = **, p<0.001 = ***, p <0.0001 = ****. |  |  |  |  |

**Figure 4:**
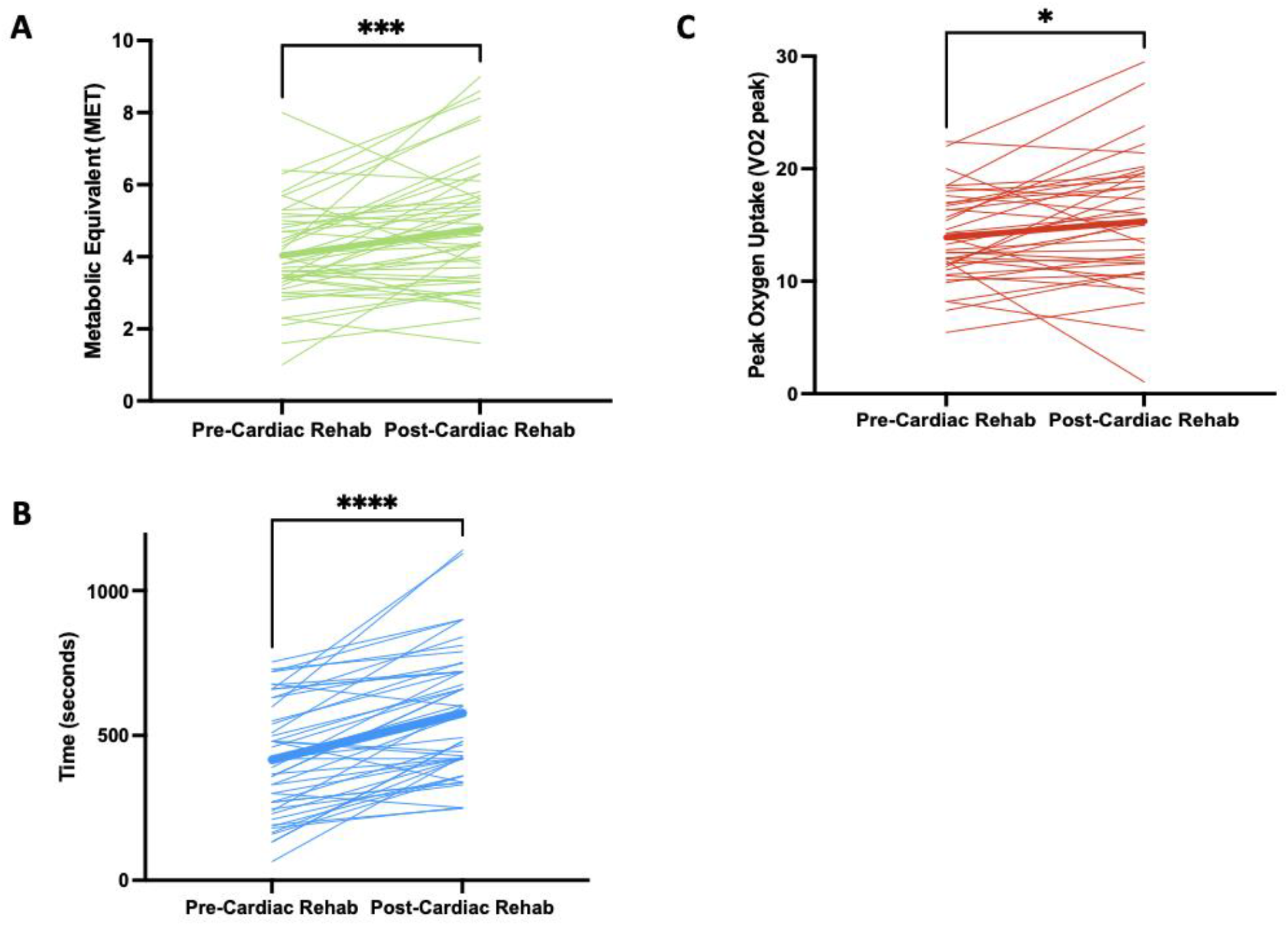
A. Metabolic equivalent task (MET) B. Exercise time C. Peak oxygen uptake before and after cardiac rehabilitation for patients with heart transplantation or left ventricular assist device implantation who completed the program. Bold lines indicate mean pre and post values. Paired-sample t tests assessed difference in pre and post cardiac rehabilitation values, * <0.05, ** <0.01, *** <0.001, **** <0.0001.

## Discussion

To our knowledge, this is the largest study to date to report CR adherence and risk factors for cessation among either heart transplant or LVAD recipients. There were several key findings. First, we found a bimodal distribution of CR attendance; nearly half of patients attended all 36 sessions, but a considerable proportion completed fewer than 24 sessions. Medical reasons were most commonly cited as cause for termination prior to 36 sessions. Second, patients who did not complete all 36 CR sessions were more likely to have experienced post-operative complications as well as post-operative major bleeding following their heart transplantation or LVAD implantation compared with those who completed CR, even after adjustment for demographics and comorbidities. Third, among patients who did complete all 36 CR sessions with pre-and post-exercise testing, there were significant improvements in exercise measures. This underscores that CR can be successfully implemented to improve functional status, even in this medically complicated population.

To our knowledge there are limited prior data on CR completeness of attendance among the heart transplant or LVAD population. Among the general HF population, the largest study conducted to date was HF-ACTION which randomized 2331 participants to usual care plus aerobic exercise training, consisting of 36 supervised sessions followed by home-based training, versus usual care alone. Sixty four percent (736 of 1159) of HF-ACTION participants in the exercise arm completed all 36 sessions.^26^ The rate of complete attendance in our sample was lower although our findings were based in an observational (rather than clinical trial) setting, and in a population that is much more medically complex. Early termination (defined as completion of fewer than 12 CR sessions) was lower in our sample than in a more general HF sample from the same database published previously (22.6% vs. 26.4%),^27^ and a higher proportion completed all 36 sessions which may reflect the expectations patients with heart transplant of LVAD have in regards to follow-up, as eligibility for these treatments is restricted to carefully selected candidates who are likely to be adherent.^28^

We also found that patients who experienced post-operative complications at the time of index hospitalization were significantly less likely to complete all 36 sessions, and most participants who did not complete all sessions (71.6%) cited medical reasons, suggesting the sequelae of these complications had long-term impacts. This differs from a separate analysis of 526 patients by Sanderson et al. where 63% participants who were unable to complete CR cited personal (and not medical) reasons.^29^ Our population had higher medical complexity than most other patients undergoing CR, which may account for this observed difference.

Notably, among our subsample of patients who completed CR, there were significant improvements in exercise time and METs as well as an increase in peak VO_2_, underscoring the effectiveness of CR in this population. This has been documented in prior studies: in a 2017 meta-analysis of randomized controlled trials that compared exercise-based rehabilitation to no exercise in a cumulative total of 300 heart transplant patients, CR significantly improved peak VO_2_ in all trials with a pooled improvement of one MET.^30^ Similarly, among six trials of 183 LVAD patients subjected to exercise-based CR or standard therapy, CR improved peak VO_2_ in all trials.^31^

There are several limitations to our study. First, our cohort only included patients who attended at least one CR session, and we were therefore unable to compare outcomes in attendees versus non-attendees. Further, we used a convenience sample that was not powered *a priori* to test for differences between groups in characteristics associated with CR cessation. Third, reasons for cessation from CR were limited to clinical documentation, and we did not directly interview patients given the retrospective nature of our database. We believe that further studies using qualitative methods may better shed light on reasons for variable attendance in this population.

In summary, we found a bimodal distribution of CR attendance among heart transplant and LVAD recipients. Postoperative complications were associated with failure to attend all 36 sessions, demonstrating a population at-risk of limited CR participation. Medical reasons for cessation were more common in our advanced heart failure sample. Finally, among those with complete data, CR was associated with significant improvements in exercise measures, underscoring the benefits of this treatment modality as also reported in prior studies.

## Data Availability

The data that support the findings of this study are available from the corresponding author, SKS, upon reasonable request.

## Acknowledgments

Dr. Dodson’s effort is supported by a mid-career mentoring award (K24AG080025) from the NIH/NIA.

